# Predicting involuntary admission following inpatient psychiatric treatment using machine learning trained on electronic health record data

**DOI:** 10.1101/2024.04.11.24305658

**Authors:** Erik Perfalk, Jakob Grøhn Damgaard, Martin Bernstorff, Lasse Hansen, Andreas Aalkjær Danielsen, Søren Dinesen Østergaard

**Affiliations:** Department of Affective Disorders, Aarhus University Hospital – Psychiatry, Aarhus, Denmark; Department of Clinical Medicine, Aarhus University, Aarhus, Denmark

## Abstract

**Background:** Involuntary admissions to psychiatric hospitals are on the rise. If patients at elevated risk of involuntary admission could be identified, prevention may be possible.

**Objectives:** To develop and validate a prediction model for involuntary admission of patients receiving care within a psychiatric service system using machine learning trained on routine clinical data from electronic health records (EHRs).

**Methods:** EHR data from all adult patients who had been in contact with the Psychiatric Services of the Central Denmark Region between 2013 and 2021 were retrieved. We derived 694 patient predictors (covering e.g., diagnoses, medication, and coercive measures) and 1,134 predictors from free text using term frequency - inverse document frequency and sentence transformers. At every voluntary inpatient discharge (prediction time), without an involuntary admission in the two years prior, we predicted involuntary admission 180 days ahead. XGBoost and Elastic Net regularized logistic regression models were trained on 85% of the dataset. The best performing model was tested on the remaining 15% of the data.

**Results:** The model was trained on 50,634 voluntary inpatient discharges among 17,968 unique patients. The cohort comprised 1,672 voluntary inpatient discharges followed by an involuntary admission. The XGBoost model performed best in the training phase and obtained an area under the receiver operating curve of 0.84 in the test phase.

**Conclusion:** A machine learning model using routine clinical EHR data can accurately predict involuntary admission. If implemented as a clinical decision support tool, this model may guide interventions aimed at reducing the risk of involuntary admission.

## Introduction

The incidence of involuntary admissions is on the rise worldwide.^1^ Involuntary admissions are used when patients are in urgent need of psychiatric inpatient treatment, but are too ill (typically psychotic) to consent.^2^ Involuntary admission can be traumatic for patients and are costly for society.^3^ Therefore, various interventions to reduce the need for involuntary admissions have been investigated.^4^ To ensure cost-effectiveness, these interventions should preferably target patients at high risk of involuntary admission. However, such individual risk assessments are complex.

Several risk factors for involuntary admission have been identified in large patient populations.^5^ However, assessing risk at the level of the individual patient is challenging due to potential interactions between risk factors, waxing and waning of risk factors, and irregular/noisy clinical data on risk factors.^6^ Recently, however, machine learning methods have been demonstrated to handle this level of complexity well. Unlike standard statistical analyses, machine learning inherently accommodate complex interactions and idiosyncrasies, and also handles large amounts of predictors and temporal dependencies within the data.^7,8^

We are aware of two prior machine learning studies having examined involuntary admission via routine clinical data.^9,10^ Both, however, fail to construct a relevant prediction task as they do not issue predictions, which is a prerequisite for clinical relevance, but merely utilize machine learning methods for identification of risk factors for involuntary admission. Additionally, both studies only consider patients with complete data in their primary analysis, which could potentially decrease the generalisability as data from real-world practice, are typically not missing at random.^6^ We have previously shown that a machine learning model trained on routine clinical data from electronic health records (EHRs) can accurately predict mechanical restraint^11^ and are currently in the process of implementing a decision support (risk reduction) tool based on this model in clinical practice. To our knowledge, no studies have used machine learning to predict involuntary admissions at the level of the individual patient using EHR data. Therefore, the aim of this study was to fill this gap in the literature.

## Methods

An illustration of the methods used in this study is shown in Figure 1.

**Figure 1.**
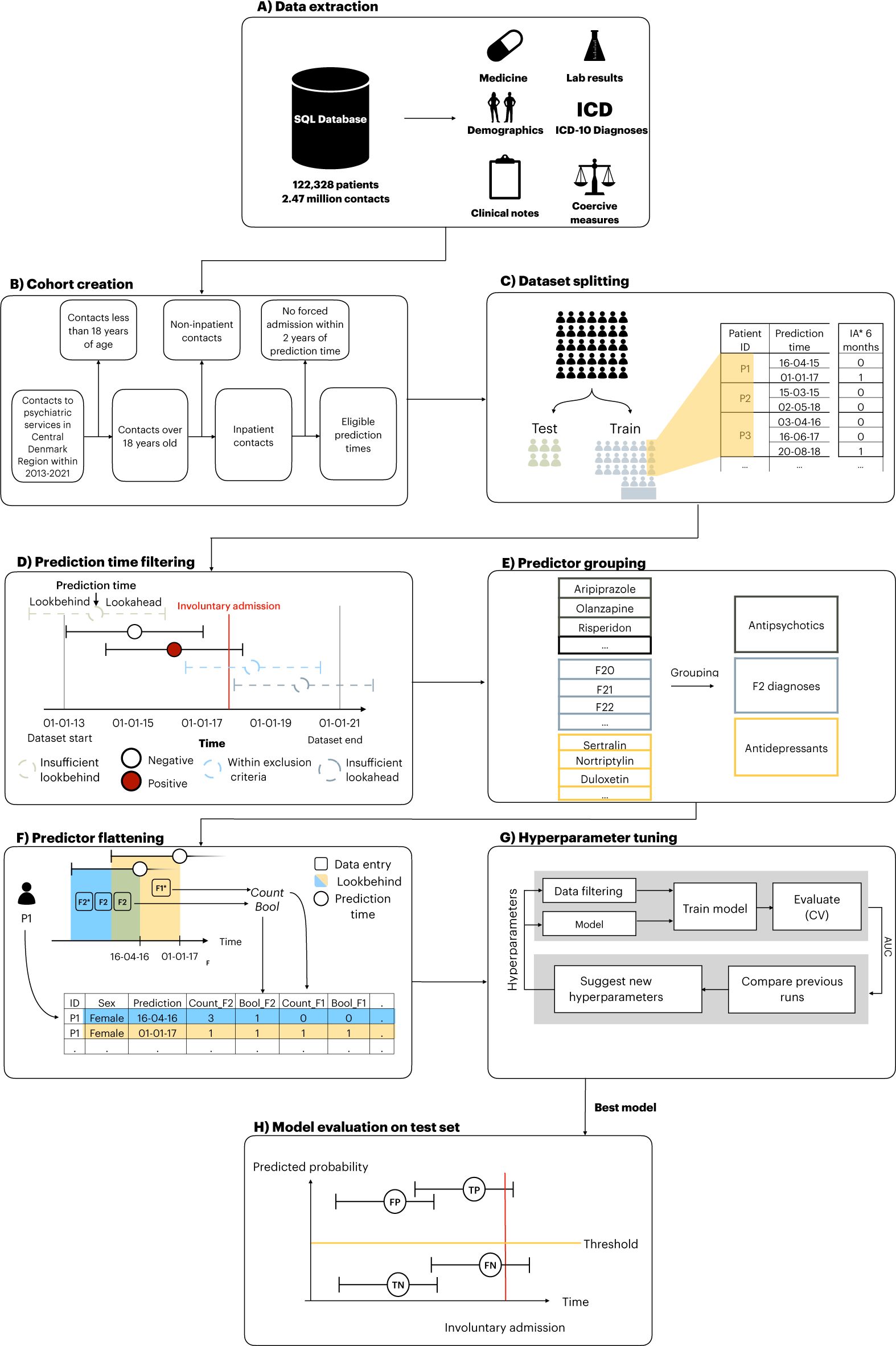
Extraction of data and outcome, dataset splitting, prediction time filtering, specification of predictors and flattening, model training, testing and evaluation. Figure 1 subtext: Figure was modified to this project based on Bernstorff et al^1^. IA: Involuntary admissions. F1 & F2: ICD-10 diagnoses within the group of diagnoses included in F1 chapter and F2 chapter. CV: Cross-validation. TP: True positive. FP: False positive. TN: True negative. FN: False negative.

### Reporting guidelines

The reporting adhered to all items considered “essential for inclusion” by at least 50% of the respondents in the first Delphi round for the Transparent Reporting of multivariable prediction models for Individual Prognosis Or Diagnosis with Artificial Intelligence (TRIPOD-AI). ^12^

### Data source

The study is based on data from the PSYchiatric Clinical Outcome Prediction (PSYCOP) cohort, encompassing routine clinical EHR data from all individuals with at least one contact to the Psychiatric Services of the Central Denmark Region in the period from January 1, 2011 to November 22, 2021.^13^ The dataset includes records from all service contacts to the public hospitals in the Central Denmark Region (both psychiatric and general hospitals). A service contact can be either an inpatient admission, outpatient visit, home visit or consultation by phone, and each is labelled with a timestamp and diagnosis. Due to the universal healthcare system in Denmark, the large majority of hospital contacts are to public hospitals (there are no private psychiatric hospitals in Denmark) and, thus, covered by these data. Importantly, the dataset also includes blood samples from general practitioners as they are analyzed at public hospitals and, as a result, are included in this dataset. ^14^

### Data extraction

All EHR data from patients with at least one contact with the Psychiatric Services of the Central Denmark Region in the period from 2013 to 2021 were extracted (Figure 1A). To ensure the feasibility of subsequent implementation of a predictive machine learning model potentially developed in this study, only data collected routinely as part of standard clinical practice and recorded in the EHR system were used (i.e., there was no data collection for the purpose of this study).^13^

### Cohort definition

Figure 1B, illustrates the cohort definition. The cohort consisted of all adult patients with at least one contact to the Psychiatric Services of the Central Denmark Region in the time period from 2011-2021. Data prior to 2013 were dropped due to data instability, primarily due to the gradual implementation of a new EHR-system in 2011.^15,16^ However, data on involuntary admissions from 2012 were used to establish incidence of involuntary admissions since these data were registered via an alternative digital system and, therefore, unaffected by the implementation of the new EHR-system.^17^

### Dataset splitting

The data was randomly split into a training (85%) and a test (15%) set by sampling unique patients, stratified by whether they had an involuntary admission within the follow-up (see Figure 1C). This ensured a balanced proportion of patients with involuntary admission in the training- and test set and prevented data leakage as no patient could occur in both datasets. The test set was not examined until the final stage of model evaluation, where no additional changes were made to the model.

### Prediction times and exclusion criteria

Prediction times were defined as the last day of a voluntary psychiatric admission. A prediction at this time point would enable outpatient clinics to initiate targeted intervention/monitoring to reduce the risk of involuntary admission. Additionally, an exclusion criterion stipulating that patients should not have had an involuntary admission in the 2 years prior the prediction time was implemented. This prevented predictions in cases where clinicians were already aware of the patient’s risk of involuntary admission, thus proactively reducing the risk of alert fatigue. Additionally, if a prediction time did not have a long enough lookbehind-(for predictors) or lookahead window (for outcomes), that prediction time was dropped. (Figure 1D). For definition of lookbehind-and lookahead windows, see the following two paragraphs.

### Outcome definition and lookahead window

The outcome was defined as the start of an involuntary admission. The lookahead window (the period following the prediction time in which the outcome could occur) was 180 days. Hence, all prediction times for which an involuntary admission occurred within 180 days were deemed to be positive outcomes (Figure 1D).

### Predictor engineering and lookbehind window

A full list of the predictors (a total of 1828) and their definitions is available in Supplementary Table 1. The predictors were chosen based on the literature on risk factors for involuntary admissions^5^ supplemented with clinical domain knowledge. All predictors were engineered with four different fixed lookbehind windows: the 10, 30, 180 and 365 days leading up to a prediction time, respectively. Additionally, different predictor aggregation methods (mean, max, bool, etc.) were employed using the timeseriesflattener v2.0.1 package (Figure 1F). ^18^ If a predictor was not present in the lookbehind period from a prediction time, it was labelled as “missing”. However, these instances do not indicate missing values in the conventional sense, as they stem from a genuine lack of data, rather than, e.g., a missed visit in a clinical trial. This absence reflects real-world clinical practice, and, therefore, patients with such missing data should not be excluded, as it aligns with the available data for potential implementation.

The predictors can be grouped into nine strata: age and sex, hospital contacts, psychiatric diagnoses, medications, lab results, coercive measures, psychometric rating scales, suicide risk assessment, and free text predictors from EHR clinical notes (extracted via natural language processing). Specifically, hospital contacts included both inpatient and outpatient contacts with linked diagnoses. Diagnoses included all psychiatric subchapters (F0-F9) from the International Classification of Disease (ICD-10) ^18^ with specific predictors for schizophrenia (F20), bipolar disorder (F30-F31) and cluster b-personality disorders (F60.2-F60.4 (dissocial-, borderline-and histrionic personality disorder). Medication predictors were based on structured Anatomical Therapeutic Chemical (ATC) classification system codes ^19^and grouped as follows (figure 1E): antipsychotics, first generation antipsychotics, second generation antipsychotics, depot antipsychotics, antidepressants, anxiolytics, hypnotics/sedatives, stimulants, analgesics, and drugs for alcohol abstinence/opioid dependence. Finally, lithium, clozapine, and olanzapine were included as individual predictors. Predictors based on laboratory tests included plasma levels of antipsychotics, antidepressants, paracetamol, and ethanol. Coercive measures included involuntary medication, manual restraint, chemical restraint, and mechanical (belt) restraint. Scores from psychometric rating scales included the Brøset violence checklist,^20^, the 17-item Hamilton depression rating scale (HAM-D17)^21^ and a simplified version of the Bech Rafaelsen mania rating scale (MAS-M).^22^ Data on suicide risk assessment was based on a scoring system used in the Central Denmark Region with the following risk levels: 1 (no increased risk), 2 (increased risk), and 3 (acutely increased risk).

Predictors from free text stemmed from the subset of EHR clinical note types deemed to be most informative and stable over time, e.g., “Subjective Mental State” and “Current Objective Mental State” (for the full list of clinical note types, see Supplementary Table 2).^15^ Two different algorithms were applied to create predictors from the free text: term frequency–inverse document frequency (TF-IDF)^23^ and sentence transformers^24^. For the TF-IDF model, the unstructured free text was first preprocessed by lower-casing all words and removing stop words and symbols. Subsequently, the model generated all uni- and bi-grams. Secondly, top 10% by document frequency were removed (due to assumed low predictive value). Lastly, the top 750 uni- or bigrams were included in the model. For each patient, all clinical notes within the 180 days lookbehind prior to a prediction time were concatenated into a single document from which the TF-IDF predictors were constructed. A pre-trained multilingual sentence transformer model^24^ was applied to extract sentence embeddings (Model: “paraphrase-multilingual-MiniLM-L12-v2”). This model is bound by a maximum input sequence length of 512 tokens. For each patient, the first 512 tokens from each clinical note within the 180 days lookbehind prior to a prediction time were extracted and input to the model, yielding a contextualized embedding of the text with 384 dimensions. Subsequently, the embeddings from each note within the lookbehind window were averaged to obtain a single aggregated embedding, which was included as a predictor in the model.

### Hyperparameter tuning and model training

Two types of machine learning models were trained: XGBoost and elastic net regularized logistic regression (using scikit learn version 1.2.1).^23^ XGBoost was chosen because gradient boosting techniques typically excel in predictive accuracy for structured data, offer rapid training, and intrinsically handle missing values.^25,26^ Elastic net regularized logistic regression served as a benchmark model.^27,28^ 5-fold stratified cross-validation was adopted for training with no patient occurring in more than one fold. Fine-tuning of hyperparameters (see Supplementary Table 3 for details) was performed to optimize the area under the receiver operating characteristic curve (AUROC) through the tree-structured parzen estimator method in Optuna v2.10.1.33 (see Figure 1G).^29^ All analyses were performed using Python (version 3.10.9).

### Model evaluation on test data

The model which achieved the highest AUROC following cross-validation on the training set was evaluated on the test set (see Figure 1H). Apart from AUROC, we also calculated the sensitivity, specificity, positive (PPV), and negative predictive values (NPV) at predicted positive rates (PPR) of 1%, 2%, 3%, 4%, 5%, 10%, 20% and 50%, respectively. The predicted positive rate is the proportion of all prediction times that are marked as positive. To test the robustness of the best performing model, its performance was examined across sex-, age-, months since first visit-, month of year, and day of week strata. Furthermore, a time-to-outcome robustness analysis was conducted to assess how the model behaved at different time-to-outcome thresholds.

### Estimation of predictor importance

Predictor importance was estimated via information gain.^29^ In the case of XGBoost, the information gain of a predictor is calculated as the change in predicted probability at a given node split, averaged across all trees in the model.

### Secondary analyses of alternative model designs

As secondary analyses, we performed model training using alternative model designs. First, we removed the implemented exclusion criterion of having an involuntary admission in the two years preceding a prediction time. Second, we assessed the importance of the number of predictors, by using only subsets of the full predictor set in the model training. Specifically, three distinct predictor sets were considered (all including sex and age): Only diagnoses, only patient descriptors (all predictors except text predictors), and only text predictors. Third, models with lookahead windows of 90 and 365 days, respectively, were trained.

### Ethics

The study was approved by the Legal Office of the Central Denmark Region in accordance with the Danish Health Care Act §46, Section 2. The Danish Committee Act exempts studies based only on EHR data from ethical review board assessment (waiver for this project: 1-10-72-1-22). Handling and storage of data complied with the European Union General Data Protection Regulation. The project is registered on the list of research projects having the Central Denmark Region as data steward.

## Results

The full dataset consisted of 52,600 voluntary admissions distributed among 19,252 unique patients. A total of 1,672 of the voluntary admissions were followed by an involuntary admission within 180 days after discharge (positive outcome), distributed across 806 unique patients (an involuntary admission can be included in multiple positive outcomes as a patient can have multiple voluntary admissions (prediction times) in the 180 days prior to an involuntary admission (positive outcome)).

Table 1 lists clinical and demographic patient data for the prediction times included in the training and evaluation of the main model. The main model included predictors with a lookbehind window of up to 365 days. After filtering away all prediction times where the lookbehind or lookahead windows extended beyond the available data for a patient, a total of 50364 prediction times remained. These prediction times were distributed across 17,968 unique patients (49.4% females (training set = 49.3% and test set = 50.0%), median age = 40.2 years (training set = 40.5 and test set = 39.2)).

**Table 1.** Descriptive statistics for prediction times.

| A | Overall | Train | Test |
| --- | --- | --- | --- |
| **n** | 50364 | 43188 | 7176 |
| **Age, median [Q1,Q3]** | 40.2 [28.3,53.5] | 40.5 [28.3,53.7] | 39.2 [28.0,52.2] |
| **Age 18-20 years, n (%)** | 998 (2.0) | 861 (2.0) | 137 (1.9) |
| **Age 20-29 years, n (%)** | 12462 (24.7) | 10630 (24.6) | 1832 (25.5) |
| **Age 30-39 years, n (%)** | 10560 (21.0) | 8965 (20.8) | 1595 (22.2) |
| **Age 40-49 years, n (%)** | 9657 (19.2) | 8209 (19.0) | 1448 (20.2) |
| **Age 50-59 years, n (%)** | 8281 (16.4) | 7157 (16.6) | 1124 (15.7) |
| **Age 60-69 years, n (%)** | 4715 (9.4) | 4091 (9.5) | 624 (8.7) |
| **Age 70-79 years, n (%)** | 2568 (5.1) | 2294 (5.3) | 274 (3.8) |
| **Age 80-89 years, n (%)** | 941 (1.9) | 808 (1.9) | 133 (1.9) |
| **Age +90 years, n (%)** | 182 (0.4) | 173 (0.4) | 9 (0.1) |
| **Female, n (%)** | 25219 (50.1) | 21495 (49.8) | 3724 (51.9) |
| **F0\* Organic mental disorder, n (%)** | 2506 (5.0) | 2186 (5.1) | 320 (4.5) |
| **F1 Substance use disorders, n (%)** | 13018 (25.8) | 11164 (25.8) | 1854 (25.8) |
| **F2 Psychotic disorders, n (%)** | 17090 (33.9) | 14640 (33.9) | 2450 (34.1) |
| **F3 Affective disorders, n (%)** | 18231 (36.2) | 15556 (36.0) | 2675 (37.3) |
| **F4 Neurotic disorders, n (%)** | 14397 (28.6) | 12371 (28.6) | 2026 (28.2) |
| **F5 Eating, sleeping and sexual disorders, n (%)** | 1653 (3.3) | 1380 (3.2) | 273 (3.8) |
| **F6 Personality disorders, n (%)** | 6395 (12.7) | 5644 (13.1) | 751 (10.5) |
| **F7 Mental retardation disorders, n (%)** | 1631 (3.2) | 1402 (3.2) | 229 (3.2) |
| **F8 Disorders of psychological development, n (%)** | 2070 (4.1) | 1703 (3.9) | 367 (5.1) |
| **F9 Child and adolescent disorders, n (%)** | 5653 (11.2) | 4719 (10.9) | 934 (13.0) |

\*(F\*) indicates the ICD-10 chapter.

### Hyperparameters and model training

The cross validation on the training set for model tuning showed that XGBoost (AUROC for the primary model = 0.79) outperformed logistic regression (AUROC for the primary model = 0.78) across all model variations (see Table 2). The hyperparameters used for the best models on the different predictor sets are listed in Supplementary Table 4.

**Table 2.**
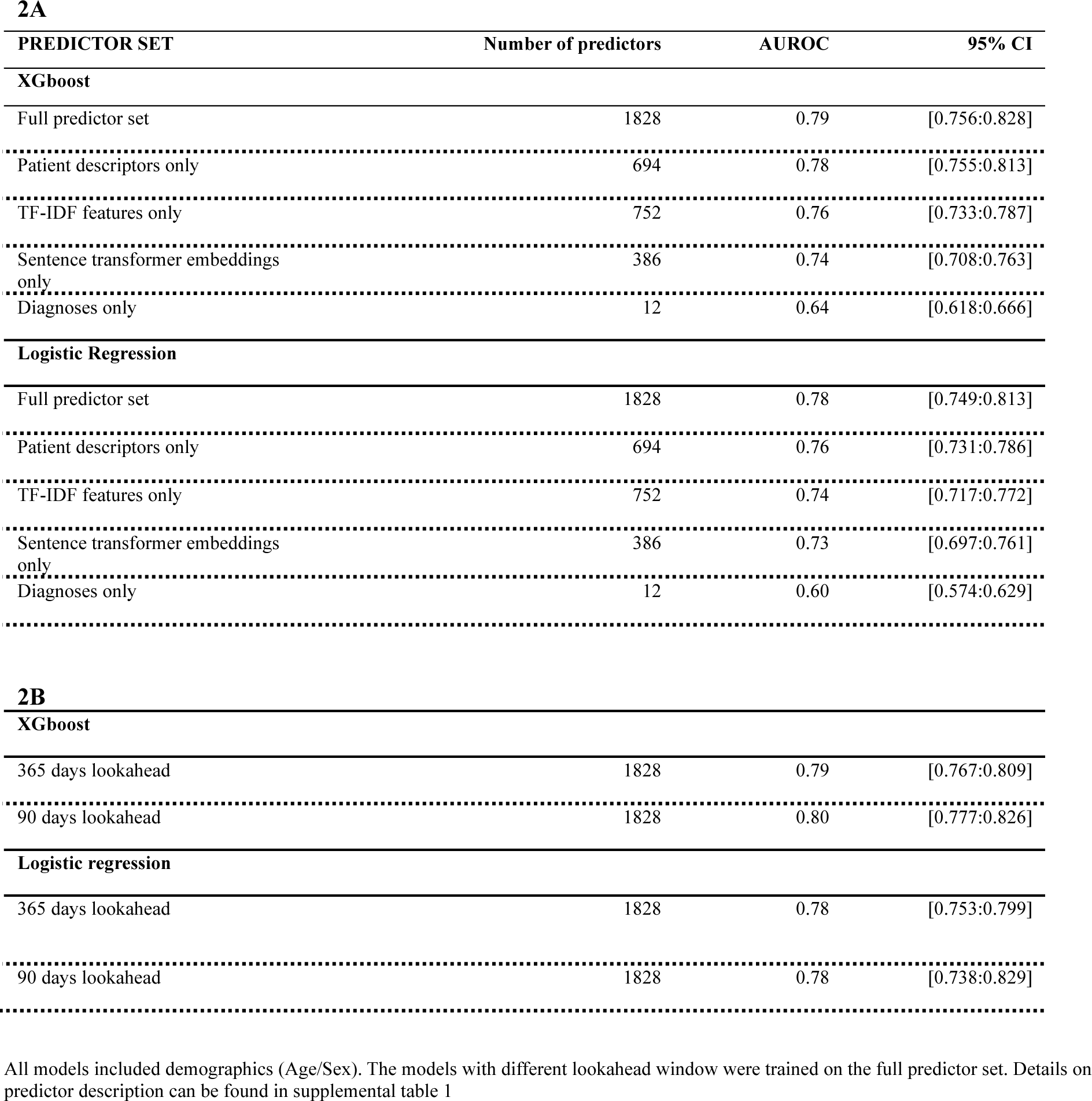
Model performance after cross-validation hyperparameter tuning for Xgboost and Logistic regression models trained on different subsets of the predictors (2A) and different lookaheads (2B)

| PREDICTOR SET | Number of predictors | AUROC | 95% CI |
| --- | --- | --- | --- |
| **XGboost** |  |  |  |
| Full predictor set | 1828 | 0.79 | [0.756:0.828] |
| Patient descriptors only | 694 | 0.78 | [0.755:0.813] |
| TF-IDF features only | 752 | 0.76 | [0.733:0.787] |
| Sentence transformer embeddings only | 386 | 0.74 | [0.708:0.763] |
| Diagnoses only | 12 | 0.64 | [0.618:0.666] |
| **Logistic Regression** |  |  |  |
| Full predictor set | 1828 | 0.78 | [0.749:0.813] |
| Patient descriptors only | 694 | 0.76 | [0.731:0.786] |
| TF-IDF features only | 752 | 0.74 | [0.717:0.772] |
| Sentence transformer embeddings only | 386 | 0.73 | [0.697:0.761] |
| Diagnoses only | 12 | 0.60 | [0.574:0.629] |
| **2B** |  |  |  |
| **XGboost** |  |  |  |
| 365 days lookahead | 1828 | 0.79 | [0.767:0.809] |
| 90 days lookahead | 1828 | 0.80 | [0.777:0.826] |
| **Logistic regression** |  |  |  |
| 365 days lookahead | 1828 | 0.78 | [0.753:0.799] |
| 90 days lookahead | 1828 | 0.78 | [0.738:0.829] |

All models included demographics (Age/Sex). The models with different lookahead window were trained on the full predictor set. Details on predictor description can be found in supplemental table 1

### Model evaluation on test data

The best performing XGBoost model identified in the training phase yielded an AUROC of 0.84 on the test set (see Figure 2A).

**Figure 2.**
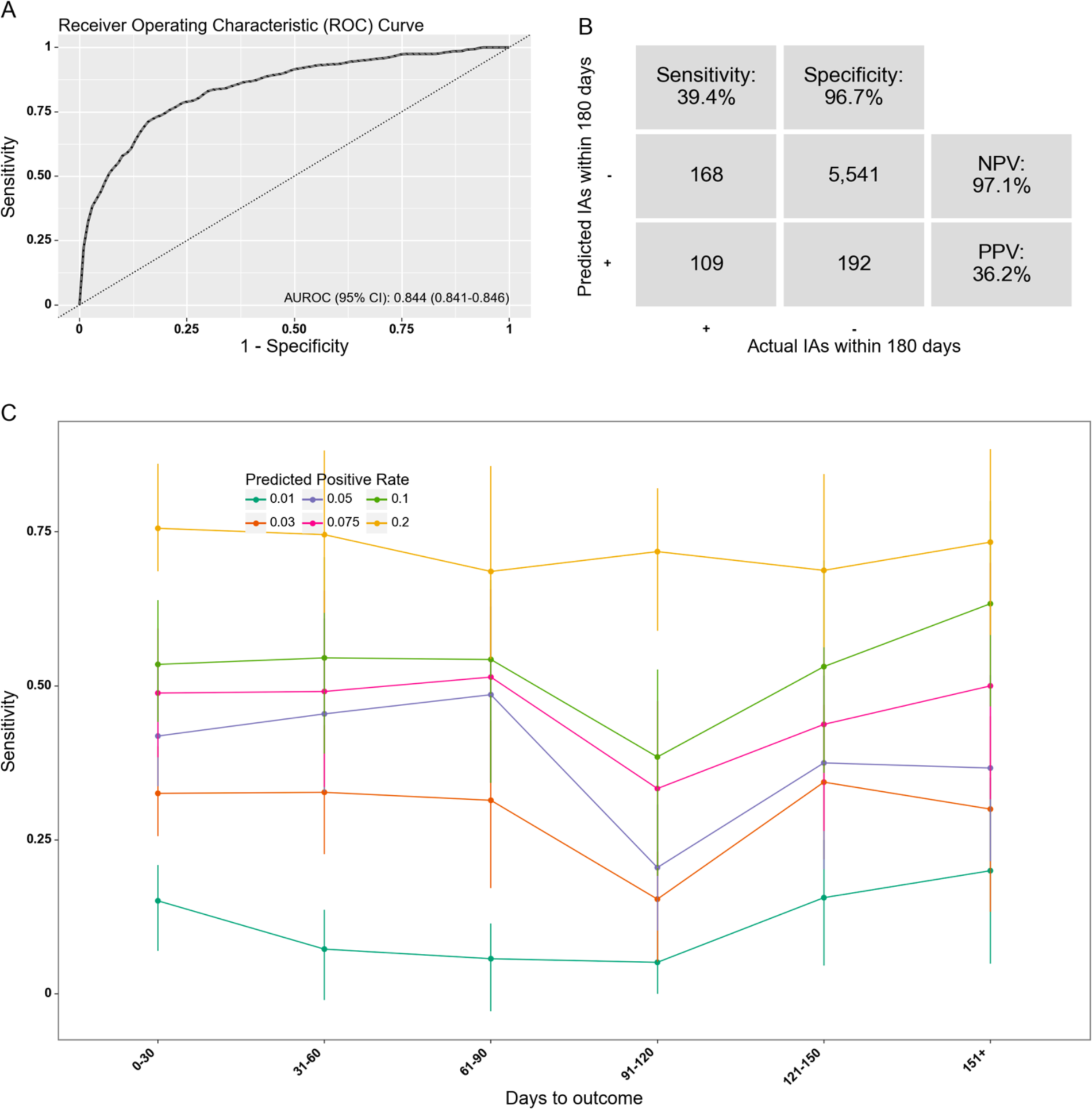
Model performance of the XGBoost model in the test set. A) Receiver operating characteristics curve. AUROC= Area under the receiver operating characteristics curve. B) Confusion matrix. PPR: Positive predictive rate. NPV: Negative predictive value. IA: Involuntary admission. The decision threshold is defined based on a predicted positive rate of 5%. C) Sensitivity (at same specificity) by months from prediction time to event, stratified by desired predicted positive rate.

Table 3 lists the performance metrics from this model on the test set based on different predicted positive rates. At a PPR of 5%, the model has a sensitivity of 39% and a positive predictive value of 36%. Thus, approximately two out of five of all true positive outcomes are correctly predicted, and for every three positive predictions, more than one prediction time is truly followed by an involuntary admission within 180 days. At this PPR, 36% of the unique involuntary admissions that underlie the positive outcomes are correctly detected (predicted positive) at least once.

**Table 3.** Performance metrics on test set for model trained on full predictor set at varying positive rates.

| Predicted positive rate | AUROC | True prevalence | PPV | NPV | Sens | Spec | FPR | FNR | Acc | TP | TN | FP | FN |
| --- | --- | --- | --- | --- | --- | --- | --- | --- | --- | --- | --- | --- | --- |
| 50.0% | 0.84 | 4.6% | 8.3% | 99.1% | 90.3% | 51.9% | 48.1% | 9.7% | 53.7% | 250 | 2,978 | 2,755 | 27 |
| 20.0% | 0.84 | 4.6% | 16.8% | 98.4% | 72.9% | 82.6% | 17.4% | 27.1% | 82.1% | 202 | 4,733 | 1,000 | 75 |
| 10.0% | 0.84 | 4.6% | 24.3% | 97.6% | 52.7% | 92.1% | 7.9% | 47.3% | 90.2% | 146 | 5,278 | 455 | 131 |
| 7.5% | 0.84 | 4.6% | 28.6% | 97.3% | 46.6% | 94.4% | 5.6% | 53.4% | 92.2% | 129 | 5,411 | 322 | 148 |
| 5.0% | 0.84 | 4.6% | 36.2% | 97.1% | 39.4% | 96.7% | 3.3% | 60.6% | 94.0% | 109 | 5,541 | 192 | 168 |
| 4.0% | 0.84 | 4.6% | 40.2% | 96.9% | 35.0% | 97.5% | 2.5% | 65.0% | 94.6% | 97 | 5,589 | 144 | 180 |
| 3.0% | 0.84 | 4.6% | 45.9% | 96.7% | 30.0% | 98.3% | 1.7% | 70.0% | 95.1% | 83 | 5,635 | 98 | 194 |
| 2.0% | 0.84 | 4.6% | 50.8% | 96.3% | 22.4% | 99.0% | 1.0% | 77.6% | 95.4% | 62 | 5,673 | 60 | 215 |
| 1.0% | 0.84 | 4.6% | 52.5% | 95.9% | 11.6% | 99.5% | 0.5% | 88.4% | 95.4% | 32 | 5,704 | 29 | 245 |
| Predicted positive rate | F1 | MCC | Total number of unique outcome events | Number of positive outcomes in test set (TP+FN) | Number of unique outcome events detected | Prop. of unique outcome events detected | Median days from first positive to outcome |  |  |  |  |  |  |
| 50.0% | 15.2% | 17.7% | 136 | 277 | 124 | 91.2% | 78 |  |  |  |  |  |  |
| 20.0% | 27.3% | 29.1% | 136 | 277 | 96 | 70.6% | 82 |  |  |  |  |  |  |
| 10.0% | 33.3% | 31.3% | 136 | 277 | 70 | 51.5% | 81 |  |  |  |  |  |  |
| 7.5% | 35.4% | 32.6% | 136 | 277 | 60 | 44.1% | 69.50 |  |  |  |  |  |  |
| 5.0% | 37.7% | 34.6% | 136 | 277 | 49 | 36.0% | 69.50 |  |  |  |  |  |  |
| 4.0% | 37.5% | 34.7% | 136 | 277 | 40 | 29.4% | 69.50 |  |  |  |  |  |  |
| 3.0% | 36.2% | 34.7% | 136 | 277 | 36 | 26.5% | 72 |  |  |  |  |  |  |
| 2.0% | 31.1% | 31.7% | 136 | 277 | 24 | 17.6% | 67 |  |  |  |  |  |  |
| 1.0% | 18.9% | 23.1% | 136 | 277 | 13 | 9.6% | 53 |  |  |  |  |  |  |

**Predicted positive rate:** The proportion of contacts predicted positive by the model. Since the model outputs a predicted probability, this is a threshold set during evaluation.
**True prevalence:** The proportion of admissions that qualified for an outcome within the lookahead window.
**AUROC** Area under the receiver operator characteristic (ROC) curve
**PPV:** Positive predictive value.
**NPV:** Negative predictive value.
**FPR:** False positive rate.
**FNR:** False negative rate.
**TP:** True positives. Numbers are based on prediction times (end of psychiatric admission).
**TN:** True negatives. Numbers are based on prediction times (end of psychiatric admission).

Figure 2C shows the sensitivity of the model for prediction times with varying time to the outcome at different PPRs. The sensitivity curves appear to remain stable as the time to outcome increases. The median time from the first positive prediction to the involuntary admission was 69.5 days.

Figure 3 shows the performance of the model across different patient characteristics and calendar time subgroups. The model appears robust across all characteristics and the minor fluctuations, such as the variation in performance between the sexes, can likely be attributed similar minor differences in sample distributions.

**Figure 3.**
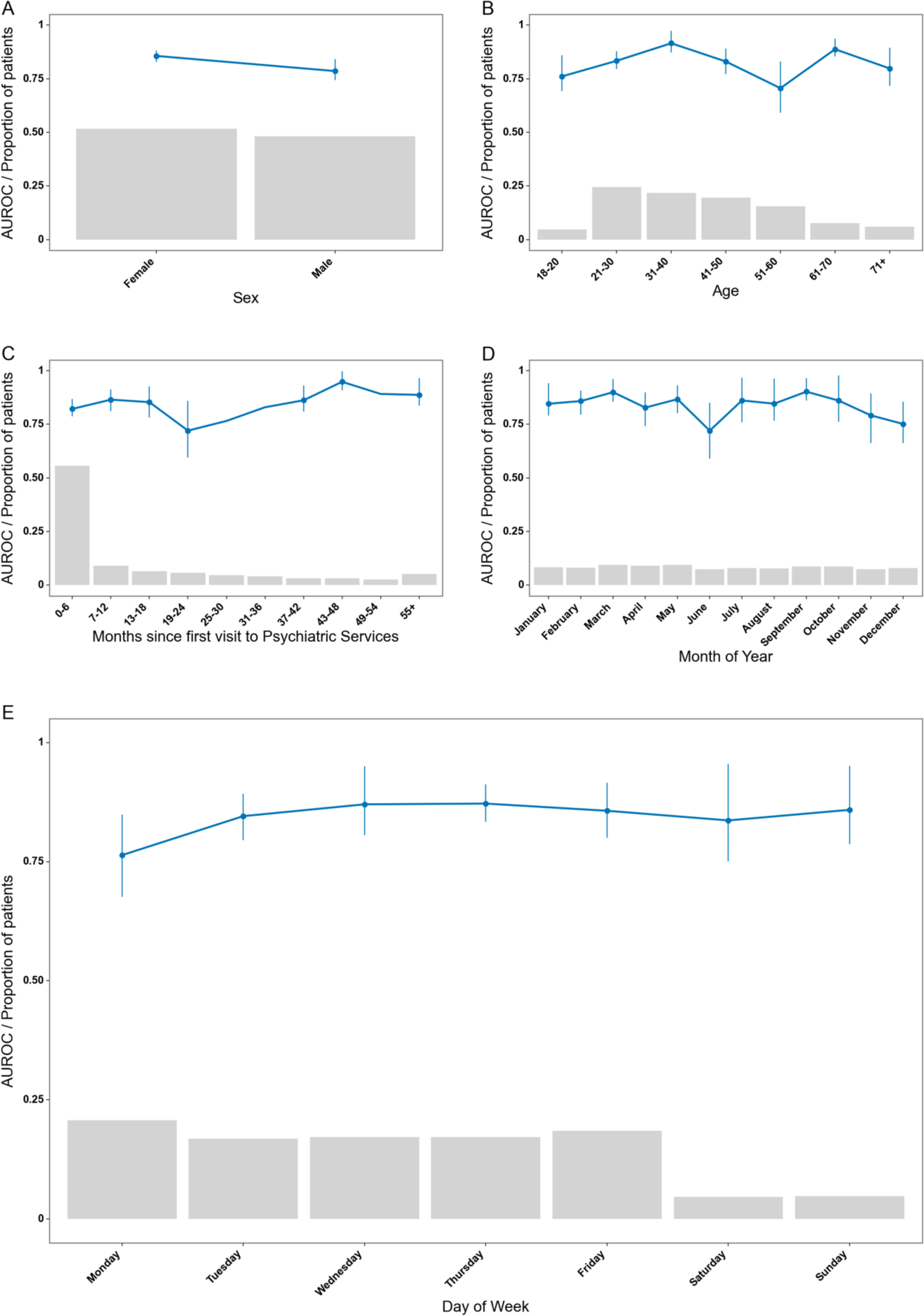
Model robustness plots stratified on demographics and time frames. Robustness of the model across different stratifications. The blue lines indicate the area under the receiver operating characteristics curve. Grey bars represent the proportion of prediction times in each bin. Error bars are 95%-confidence intervals from 100-fold bootstrap. Due to the low number in some of the bins, some bootstrapped folds contained only one class, resulting in missing error bars.

Supplementary Table 5 lists the 30 predictors with the highest information gain. Out of the 30 top predictors, 14 were text predictors – both represented by TF-IDF and sentence transformers. The TF-IDF predictors were based on the following terms from free text: “ECT”, “police”, “social psychiatric institution”, “self-harm”, “woman”. The 16 remaining predictors were distributed on the following patient descriptors: detention, coercion due to danger to self or others, lab test of plasma-paracetamol, Brøset violence checklist score, diagnosis of child and adolescent disorder/unspecified mental disorder (ICD F9-chapter), visit to a physical department, suicide risk assessment score, diagnosis of personality disorder (ICD F6-chapter).

### Secondary analyses on alternative model designs

The model which was trained without the exclusion criterion of having an involuntary admission in the 2 years preceding the prediction time yielded an AUROC of 0.90 on the cross-validated training set. Performance of the cross-validated models using different subsets of the full predictor set (Table 2A) and different lookaheads (Table 2B) are shown in Table 2. Among those trained on different subsets of predictors, the best performing model was the one trained on only patient descriptors (no text). In the models trained on different outcome lookaheads, the model with a 90 day lookahead performed better than the ones with 180 and 365 day lookaheads.

## Discussion

This study investigated if involuntary admission can be predicted using machine learning models trained on EHR data. When issuing a prediction at the discharge from a voluntary inpatient admission, based on both structured and text predictors, the best model (XGBoost) performed with an AUROC of 0.84. The model was generally stable across different patient characteristics, calendar times, and with varying times from prediction to outcome.

To our knowledge, this is the first study to develop and validate a prediction model for involuntary admission using routine clinical data from EHRs. We can, therefore, not offer a direct comparison of our results to those from other studies. However, a crude comparison to other prediction studies in psychiatry shows that our results are within the performance ranges that have previously been published.^30^ Many of these studies have, however, not been developed on routine clinical data, but rather on data collected for the purpose of the studies, which complicates clinical implementation.

On the independent test set, the prediction model performed with an AUROC above the upper boundary of the confidence interval estimated from the 5-fold cross-validation in the training phase. While this suggests that the model did not overfit on the training set, the variation in model performance might be attributable to the limited number of positive cases in the test set. Tests on larger datasets will provide further knowledge on the robustness of the model. The prediction model demonstrated relatively stable sensitivity when increasing time from prediction to outcome (up to several months), highlighting that model performance is not merely driven by prediction of cases where an involuntary admission occurs shortly after discharge from a voluntary admission. Indeed, the median time from the first positive prediction to the involuntary admission of 69.5 days is sufficient to issue a potentially preventive intervention through, e.g., advance statements/crisis plans.^4^

A series of secondary analyses were conducted to explore the impact of various model design decisions. First, the exclusion criterion stipulating that patients could not have had an involuntary admission in the two years prior to a prediction time was added to minimize the potential alert fatigue in clinicians. Specifically, this measure aimed to omit scenarios where clinicians are likely already aware of an increased risk of involuntary admission, given that prior involuntary admission is a major risk factor for subsequent involuntary admission.^5^ Indeed, this was confirmed by our results as the model trained without this exclusion criterion performed with an AUROC of 0.90 (on the cross-validated training set). This highlights the challenging balance between minimizing potential alert fatigue among clinicians and optimizing model performance for prediction models in healthcare. Second, the performance of a model trained on a limited feature set including only age, sex and diagnoses was tested, resulting in an AUROC of 0.64 (on the cross-validated training set). This demonstrates that using the full predictor set resulted in substantially better predictive performance, underlining the complexity of risk prediction at the level of the individual patient. Third, a lookahead window of 180 days was chosen for the main model as this leaves a reasonable window of opportunity for prevention of an involuntary admission. Models trained with lookahead windows of 90 and 365 days achieved an AUROC of 0.80 and 0.79, respectively, using 5-fold cross validation on the training set. This further validates the performance-wise stability of the method across different time-to-outcome intervals and justifies determining the optimal lookahead window based on clinical judgement.

With regard to the predictors driving the discriminative abilities of the model, text features comprised 14 out of the top 30 predictors in ranked information gain, showcasing the importance of including text. This might be especially true for the field of psychiatry where the condition of a patient is mainly described in natural language in the EHR rather than in structured variables., The inclusion of predictors based on TF-IDF and sentence transformer features/embeddings of the text also over all indicated an increased performance of the model. This is in line with prior results of both our own^11^ and others.^31,32^ Among predictors extracted from the free text using TF-IDF, “ECT”, “police” and “self-harm” were among the predictors with the highest predictive value, which makes intuitive sense from a clinical perspective (proxies for severity). Regarding patient descriptors, prior detention, coercion due to danger to self or others, Brøset violence checklist score, and suicide risk assessment scores were among the top predictors, which again could be proxies for the severity of the mental illness. Furthermore, a lab test of plasma-paracetamol – another top predictor - likely indicates that a patient has taken a toxic dose of paracetamol in relation to self-harm or a suicide attempt – also a manifestation of severe mental illness. Lastly, a diagnosis of personality disorder (ICD F6-chapter), which includes borderline personality disorder and is a known risk factor for involuntary admission, was also found to be one of the most informative features.^33^

The information gain estimates should, however, be interpreted with caution as XGboost includes a random factor in the model training process. Additionally, due to the structure of a decision tree model, top predictors containing mutual information can be omitted. Consequently, this may lead to only one of the mutual information predictors being included in the information gain table.^34^ The most important insight from the information gain estimates may be that the model is not informed by a few dominant predictors, but instead relies on a plethora of predictors. In line with this, the model with a limited feature set performed with an AUROC of 0.64 on the training set. This demonstrates the complexity of the outcome and aligns with our assumption that machine learning models are well suited for this prediction task.

There are limitations to this study, which should be taken into account. First, there is a limited number of outcomes (involuntary admissions) in the dataset, and the main model considered a total of 1828 predictors. If not handled properly, this could result in “curse of dimensionality”^35^ and lead to potential overfitting. To mitigate this, we employed several strategies: structured predictors were constructed based on findings from prior research and clinical domain knowledge, we used cross-validation during training, and, during hyperparameter tuning, feature selection was adopted. Finally, we used a hold-out test-set to ensure that potential overfitting during the training phase is accounted for in the evaluation. Second, the test set was not independent with regard to time or geographic location, which could have an impact on the generalizability of the model. Machine learning models inherently vary in their generalizability and reusing our model 1:1 in another hospital setting would probably result in reduced performance. However, the overall approach is likely to be generalizable and, thus, retraining the model on another EHR-dataset, while keeping the same architecture, could enable transferability.^36^ Third, despite several text predictors demonstrating high predictive value, the methods for obtaining the predictors from the free-text notes were relatively simple. In future studies, we believe it may be possible to unlock vastly more predictive value from the text by applying more advanced language models. Specifically, a future direction could involve a transformer-based model fine-tuned specifically to psychiatric clinical notes and the given prediction task.^37^ Fourth, the approach in this project is characterized by fitting a classical binary prediction framework to a task that is inherently sequential in nature. As sequential transformer-based models are gradually adapted from language modelling to the general health care domain, it is likely that such architectures may be better suited to this task and will enhance performance. The adaptation of transformer-based models to the healthcare domain is, however, still in an explorative phase, and, hence, we deem that involving such methods in this study - which was aimed at developing a model for potential clinical implementation - would be premature.

## Conclusion

A machine learning model using routine clinical data from EHRs can accurately predict involuntary admission. If implemented as a clinical decision support tool, this model may guide interventions aimed at reducing the risk of involuntary admission.

## Data Availability

The data used for this study cannot be shared according to Danish Law as it can be used to identify individual patients.

## Contributors

The study was conceptualized and designed by all authors. Funding was raised by SDØ. The data was procured by SDØ. The statistical analyses were carried out by EP and JGD. All authors contributed to the interpretation of the results. EP wrote the first draft of the manuscript, which was subsequently revised for important intellectual content by the remaining authors. All authors approved the final version of the manuscript prior to submission.

## Acknowledgments

The authors are grateful to Bettina Nørremark for data management.

