## Supplementary material for "Predicting involuntary admission following inpatient psychiatric treatment using machine learning trained on electronic health record data"

### TABLE OF CONTENTS

#### TABLES

**Supplementary Table 1: Overview over patient descriptors and text embeddings/features used for predictor engineering**

**Supplementary Table 2: EHR clinical note types used for text features/embeddings**

**Supplementary Table 3: Hyperparameter tuning options during model training**

**Supplementary Table 4: Hyperparameters from the best models during cross-validation on different predictor sets**

**Supplementary Table 5: Information gain of top 30 model predictors from main model (XGboost, full predictor set)**

### Supplementary Table 1: Overview of patient descriptors and text embeddings/features used for predictor engineering

| Group | Patient descriptor | Lookbehind (days) | Aggregation | N |
|---|---|---|---|---|
| **Static** | Sex (male/female)<br>Age (years) | - | - | 2 |
| **Hospital contacts** | All contacts<br>Psychiatric contacts<br>Somatic contacts | 10, 30, 180, 365 | Count | 8 |
| | Admissions | 10, 30, 180, 365 | Count, sum of hours | 8 |
| **Diagnoses (ICD-10)** | F0: Organic mental disorders<br>F1: Mental and behavioural disorders due to psychoactive substance use<br>F2: Schizophrenia, schizotypal, and delusional disorders<br>F20: Schizophrenia<br>F25: Schizoaffective<br>F3: Mood/affective disorders<br>F30-F31: Manic and bipolar disorders<br>F4: Neurotic, stress-related, and somatoform disorders<br>F5: Behavioural syndromes associated with physiological disturbances and physical factors<br>F6: Disorders of adult personality and behaviour<br>F60.2-60.4 Cluster B personality (dissocial-, borderline- and histrionic personality disorder)<br>F7: Mental retardation<br>F8: Disorders of psychological development<br>F9: Behavioural and emotional disorders with onset usually occurring in childhood and adolescence or unspecified mental disorder | 10, 30, 180, 365 | Boolean, count | 112 |
| **Medication** | Antipsychotics<br>1.generation<br>2. Generation<br>Olanzapine<br>Clozapine<br>Depot antipsychotics<br>Olanzapine<br>Aripiprazole<br>Risperidone<br>Paliperidone<br>Haloperidol<br>Perphenazine<br>Zuclopenthixol<br>Anxiolytics<br>Hypnotics and sedatives<br>Antidepressants<br>Lithium<br>Alcohol dependence medications<br>Opioid dependence medications<br>Nervous system stimulants<br>Analgesics | 10, 30, 180,365 | Boolean, count | 160 |
| **Coercive measures** | Detention<br>Forced detention<br>Criteria for detention: Danger to self or others | | | |
| | Compulsory treatment<br>Involuntary medication<br>Electroconvulsive therapy<br>Involuntary treatment of somatic illness<br>Physical force<br>Manual restraint<br>Mechanical restraint<br>Mechanical restraint with straps | 10, 30, 180, 365 | Boolean, count, sum of hours | 132 |
| | Chemical restraint | 10,30,180,365 | Boolean, count | 8 |
| **Psychometric rating scales** | Brøset Violence Checklist score<br>Suicide Risk Assessment<br>Hamilton-d17 score<br>Modified Bech-Rafaelsen Mania Scale score | 10, 30, 180, 365 | Mean, maximum, minimum, slope/change_per_day, variance | 80 |
| **Lab results** | Plasma-lithium<br>Plasma-clozapine<br>Plasma-olanzapine<br>Plasma-aripiprazole<br>Plasma-risperidone<br>Plasma-paliperidone<br>Plasma-haloperidol<br>Plasma-paracetamol<br>Plasma-ethanol<br>Plasma-nortriptyline<br>Plasma-clomipsramine | 10, 30, 180, 365 | Mean, maximum, minimum, latest | 176 |
| | Cancelled lab tests | 10, 30,180, 365 | Boolean, count | 8 |
| **Text features/embeddings** | Term-frequency Inverse document frequency features | 180 | All notes within 180 days from a prediction time were | 750 |

| | | | | |
|---|---|---|---|---|
| | | | concatenated to a single document and TF-IDF scores were calculated for the 750 uni- and bi-grams in the vocabulary[f]. | |
| | Sentence transformers embeddings | 180 | Embeddings, each with 384 dimensions, were generated from the first 512 tokens of each note within the 180 days window These were then averaged to create an aggregate embedding with 384 dimensions. | 384 |

### Supplementary Table 2: EHR clinical note types used for text features/embeddings

| EHR clinical note types |
|---|
| Subjective mental state |
| Subjective physical state |
| Current objective mental state |
| Current social functioning |
| Semistructured diagnostic interview |
| Observation of patient |
| Reason for contact |
| Telephone consultation note |
| Appointments |
| Consultation with a Treatment Objective |
| Conclusion/final assessment |

### Supplementary Table 3: Hyperparameter tuning options during model training

| **Preprocessing** | |
|---|---|
| Imputation method for predictors with no data | Most frequent value, mean, median or no imputation (only possible for XGBoost). |
| Scaling | z-score-normalisation or no scaling |
| Predictor selection | Chi-squared or none |
| Predictor selection percentile[a] | Between 1 and 90 (N/A). |
| Lookbehind combination (days)[b] | One of either<br>• [10,30, 180, 365]<br>• [10,30,180]<br>• [10,180]<br>• [10,180,365] |
| **Model hyperparameters** | |
| **XGBoost** | |
| N estimators | [100; 1200] |
| Alpha | [$10^{-8}$; 0.1] |
| Lambda | [$10^{-8}$; 1.0] |
| Max depth | [1; 10] |
| Learning rate | [$10^{-8}$; 1] |
| Gamma | [$10^{-8}$; $10^{-4}$] |
| Grow policy | Either depthwise or lossguide |
| **Logistic regression** | |
| Penalty solver | Elasticnet SAGA |
| C | [$10^{-5}$; 1] |
| L1 ratio | [$10^{-5}$; 1] |

[a] Feature selection rank-orders possible predictors by their chi-squared value. For e.g., the 10-th percentile, the top 10% of

features with the highest chi-squared values were kept, the rest were dropped.
b Features were constructed for lookbehinds of 10, 30, 180, 365,. Which subset to train on was established as a hyperparameter, and one of the above combinations was selected for each training run.

### Supplementary table 4: Hyperparameters from the best models during cross-validation on different predictor sets

| Predictor set | Preprocessing | | | | | | Model hyperarameters | | | | | | |
|---|---|---|---|---|---|---|---|---|---|---|---|---|---|
| | Imputation | Scaling | Predictor selection | Predictor selection percentile | Lookbehind combination | Model | N estimators | Alpha | Lambda | Max deptha | Learning rate | Gamma | Grow policy |
| Diagnoses | None | z-score normalization | None | 83 | [10,30,180, 365] | XGboost | 507 | 0.002 | $1.31e^{-05}$ | 1 | 0.082 | $1.08e^{-05}$ | depthwise |
| Patient descriptors | None | None | None | 80 | [10,180,365] | XGboost | 719 | $1.09e^{-06}$ | $1.18e^{-06}$ | 5 | 0.013 | $5.86e^{-05}$ | loss guide |
| Sentence transformer embeddings | Mean | z-score normalization | None | 33 | [10,30,180, 365] | XGboost | 808 | 0.072 | 0.044 | 5 | 0.008 | $2.91e^{-06}$ | lossguide |
| TF-IDF features | Most frequent | z-score normalization | None | 46 | [10,30,180, 365] | XGboost | 846 | $3.06e^{-08}$ | 0.461 | 3 | 0.014 | $7.19e^{-06}$ | depthwise |
| Full predictor set | None | z-score normalization | None | 13 | [10,30,180, 365] | XGboost | 210 | 0.061 | 0.396 | 2 | 0.038 | 0.0002 | depthwise |

### Supplementary Table 5: Information gain of top 30 model predictors from main model (XGboost, full predictor set)

| **LOOKBEHIND WINDOW** | AGGREGATION FUNCTION | PREDICTOR* | INFORMATION GAIN |
|---|---|---|---|
| **180** | TF-IDF | ”ECT” | 0.020 |
| **180** | Count | Detention | 0.017 |
| **365** | Boolean | Detention | 0.016 |
| **365** | Summed | coercion due to danger to self or others | 0.016 |
| **365** | Latest | Plasma paracetamol | 0.013 |
| **180** | Sum | coercion due to danger to self or others | 0.012 |
| **180** | Mean | Brøset violence checklist score | 0.012 |
| **180** | Sentence transformer | Sentence embedding 308 | 0.011 |
| **180** | TF-IDF | ”police” | 0.011 |

| | | | |
|---|---|---|---|
| **365** | Maximum | Brøset violence checklist score | 0.011 |
| **180** | Sentence transformer | Sentence embedding 294 | 0.011 |
| **365** | Boolean | Diagnosis of F9 disorders | 0.011 |
| **180** | Sentence tranformer | Sentence embedding 344 | 0.011 |
| **180** | Count | Visits due to a physical disorder | 0.011 |
| **180** | Maximum | Suicide risk assessment score | 0.011 |
| **180** | TF-IDF | ”social psychiatric institution” | 0.011 |
| **180** | Sentence transformer | Sentence embedding 122 | 0.011 |
| **10** | Boolean | Detention | 0.011 |
| **180** | TF-IDF | ”self-harm” | 0.010 |
| **180** | Sentence transformer | Sentence embedding 79 | 0.010 |
| **365** | Minimum | Plasma paracetamol | 0.009 |
| **180** | TF-IDF | ”the week” | 0.009 |
| **180** | Sentence transformer | Sentence embedding 276 | 0.009 |
| **180** | Sentence transformer | Sentence embedding 219 | 0.009 |
| **180** | TF-IDF | ”woman” | 0.009 |
| **365** | Count | Diagnosis of F6 disorders | 0.009 |
| **365** | Mean | Suicide risk assessment score | 0.009 |
| **365** | Count | coercion due to danger to self or others | 0.009 |
| **365** | Mean | Brøset violence checklist score | 0.009 |
| **180** | Sentence transformer | Sentence embedding 255 | 0.009 |

*TF-IDF predictor labels are translated from Danish.
